# COVID-19 Twitter-based analysis reveals differential concerns across areas with socioeconomic disparities

**DOI:** 10.1101/2020.11.18.20233973

**Authors:** Yihua Su, Aarthi Venkat, Yadush Yadav, Lisa B. Puglisi, Samah J. Fodeh

**Author notes:** Co-first authors, contributed equally to this work (Yihua Su, Aarthi Venkat). **Corresponding Author Information:** Samah J. Fodeh, 300 George Street PO Box 208009, New Haven, CT 06520.

## Abstract

**Objective:** We sought to understand how U.S. residents responded to COVID-19 as it emerged, and the extent to which spatial-temporal factors impacted response.

**Materials and Methods:** We mined and reverse-geocoded 269,556 coronavirus-related social media postings on Twitter from January 23^rd^ to March 25^th^, 2020. We then ranked tweets based on the socioeconomic status of the county they originated from using the Area Deprivation Index (ADI); that we also used to identify areas with high initial disease counts (“hotspots”). We applied topic modeling on the tweets to identify chief concerns and determine their evolution over time. We also investigated how topic proportions varied based on ADI and between hotspots and non-hotspots.

**Results:** We identified 45 topics, which shifted from early-outbreak-related content in January, to the presidential election and governmental response in February, to lifestyle changes in March. Highly resourced areas (low ADI) were concerned with stocks, social distancing, and national-level policies, while high ADI areas shared content with negative expression, prayers, and discussion of the CARES Act economic relief package. Within hotspots, these differences stand, with the addition of increased discussion regarding employment in high ADI versus low ADI hotspots.

**Discussion:** Topic modeling captures the major concerns in COVID-19-related discussion on a social media platform in the early months of the pandemic. Our study extends previous studies that utilized topic modeling on COVID-19 related tweets and linked the identified topics to socioeconomic status using ADI. Comparisons between low and high ADI areas indicate differential Twitter discussions, corresponding to greater concern with economic hardship and impacts of the pandemic in less resourced communities, and less focus on general public health messaging.

**Conclusion:** This work demonstrates a novel framework for assessing differential topics of conversation correlating to income, education, and housing disparities. This, with integration of COVID-19 hotspots, offers improved analysis of crisis response on Twitter. Such insight is critical for informed public health messaging campaigns in future waves of the pandemic, which should focus in part specifically on the interests of those who are most vulnerable in the lowest resourced health settings.

## INTRODUCTION

The novel severe acute respiratory syndrome coronavirus, SARS-CoV-2, which causes the disease COVID-19, has led to a global pandemic over the course of just a few months. With no specific treatment for the disease and fears of the burden of illness overwhelming health systems, the primary public health focus has been on disease mitigation strategies [1-3]. These strategies have introduced new concepts to the general public, such as social distancing and recommendations for routine masking. These mitigation efforts along with others, including travel bans, shelter-in-place orders and school closures, were anticipated to negatively affect many sectors of the U.S. economy, and they have drastically changed the quotidian lives of most Americans. Given marked community-level socioeconomic disparities and segregation in the U.S. that predated COVID-19, the impacts of these measures were likely to have disparate uptake by and impact on Americans depending on where they live [4].

With the expansive geography of the United States (U.S.), and modern-day travel patterns, the disease initially was focused in a few cities, and these so-called “hotspots” were a primary focus of much of the initial media coverage [5]. Despite this focus, other COVID-19 hotspots with large marginalized populations later emerged [6,7], highlighting the importance of understanding differential reactions to the crisis, as this could be critical for shaping future public health communication and allocation of health resources.

Social media has been a prominent venue for personal and public health communication, both in previous public health crises and now too with COVID-19. Pre-COVID-19, social media research in the context of health was primarily focused on examining the patient experience [8-12]. Comments and reviews on Twitter were used to measure healthcare quality [10] and monitor health status of patients along with sentiment level [12]. Twitter, specifically, has the advantage of short, real-time content availability with quick access to a network of similar discussions through hashtags. It has been beneficial in various research areas for its openness and availability [13]. It has also been useful in understanding social networks, public health messaging, and forecasting spread [14-17]. Twitter played an important role in Ebola outbreak surveillance by detecting the epidemic nearly a week before its first case [15]. Influenza infection rate [16] and ZIKV case number [17] predictions, learned from the tweet count pattern of disease-related tweets, were also proven successful.

During COVID-19, Twitter has been used to assess mitigation strategies such as social distancing [18] [19], capture self-reported symptoms of COVID-19 [20], Twitter, however, has not, to our knowledge, been used as a tool to identify trends in public responses to a health crises at the local level, while factoring in socioeconomic status. Using public health communication to mitigate health disparities is not a novel concept [21], and is in line with future directions laid out in the National Institute on Minority Health and Health Disparities 2019 research framework [22], but the science on implementation of this approach is underdeveloped and is an area of active research. In this study, we sought to assess a novel approach to use Twitter to understand how COVID-19 related concerns differed by area socioeconomic status in the initial phase of the COVID-19 pandemic in the United States.

## MATERIALS AND METHODS

### Twitter Dataset

The dataset we used for this analysis is composed of Twitter entries (tweets) in English posted by users in the United States from January 23^rd^ to March 25^th^ 2020. We mined the tweets with a Standard Search API using the keywords ‘coronavirus’, ‘corona virus’, ‘corona’, ‘covid’, ‘covid-19’, ‘covid 19’ and ‘covid19’. For each tweet, we obtained standard attributes: unique de-identified user ID, time of tweet, text of the tweet and four geographic coordinates (latitude and longitude) delineating the bounding box [23] from which the tweet was posted. For privacy reasons, Twitter does not provide the exact location that tweets were posted from. **Figure 1** demonstrates the overall workflow which will be further detailed in the following sections.

**Figure 1.**
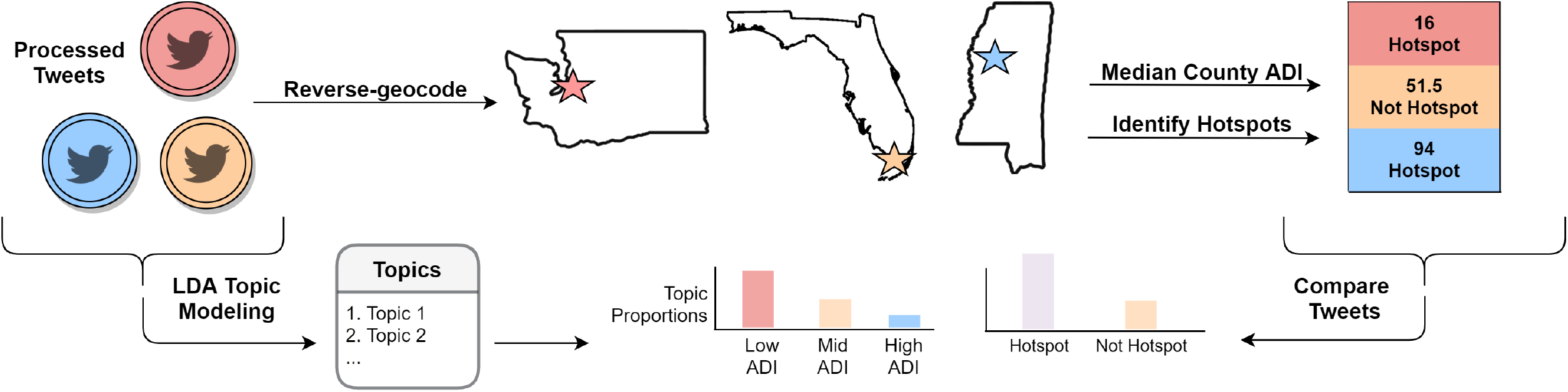
Data Integration and Analysis Workflow.

### Preprocessing of Tweets

We pre-processed through the removal of punctuation marks, numbers, emojis, URLs, stop words, and end of line characters. We shortened the remaining words to the root using the stemmer package provided by the NLTK toolkit [24]. We removed tweets that were with missing or invalid data such as those without a month or date of entry, valid user ID entry or valid stemmed tweet text. Finally, we filtered out tweets containing only words that occurred in less than 20 documents or more than 50% of all documents (of which only “coronavirus” was excluded) in order to achieve better topic models. This is a common approach [25, 26], used to avoid spurious associations by excluding words based on term distribution.

### Reverse Geocodes of Tweets

We employed GeoPy [27] to reverse geocode the coordinates and output the county and state name of each tweet. As the bounding box provides enough information to confidently geotag the tweet at the county resolution, we used the midpoint of the rectangle of latitude and longitude coordinates of each tweet as the effective location. This location was then linked to a five-digit FIPS code, a code designed to uniquely identify counties and states in the U.S., to determine the location of tweets at the county level. We followed a similar approach in our previous work [19] to map tweets to the county level.

### Area Deprivation Index (ADI) Designation

We leveraged ADI from The Neighborhood Atlas [28], a location-based socioeconomic index at the census block group level which incorporates income, education, employment and housing data and has been used to inform health delivery and policy. ADI scores range from 0 to 100, where 0 corresponds to low deprivation and 100 corresponds to high deprivation. We mapped the location of each tweet, derived from the reverse geocoding tweets process, to the median ADI score of all the census block groups within the county using its FIPS code. Counties were considered “low”, “mid”, or “high” ADI based on the ADI distribution of the unique counties represented in the dataset. Low ADI designation was assigned to counties from the lowest quintile of the distribution, and high ADI designation was assigned to counties from the highest quintile of the distribution as has been done with other studies using ADI [29, 30].

### Hotspot Identification

We defined hotspots in January and February as areas with any cases of COVID-19 because there were few U.S. cases in these months and they were concentrated (also as published by the New York Times [31]). For analyzing hotspots in March, we leveraged the curated resource The U.S. COVID-19 Atlas [32], defining a tweet as from a hotspot if the county was listed among the published population-adjusted hotspots.

### Topic Modeling

We performed topic modeling of the tweets using a Latent Dirichlet Allocation (LDA) approach [33]. LDA is an unsupervised approach and has been proven successful in modeling topics in tweets [34]. We leveraged LDA from the MALLET package [35] to detect topics from COVID-related tweets. To determine the optimal number of topics, we compared topics by their coherence scores, which act as a proxy for interpretability by measuring the degree of semantic similarity between top words in the topic [36]. We used the topic-word distribution to annotate topics. We first ranked words of a topic and then assigned the underlying theme.

### Spatiotemporal Analysis

We leveraged the document-topic probability distribution for this analysis. We compared topic prevalence over time, across low and high ADI areas, between hotspots and non-hotspots areas, and within hotspots between low ADI and high ADI areas.

#### Temporal analysis of topic prevalence

To study topic evolution and how the public reactions to COVID-19 varied temporally, we averaged the topic distributions of all tweets for each month. We then compared the average scores of all topics over time. For selected topics, we plotted out the daily topic dynamic to demonstrate how the daily average topic distribution changed.

#### Spatial analysis of topic prevalence

To compare the dominant topics in counties of low versus high ADI designation, we computed the log odds ratios of dominant topics in both groups. We first identified the dominant topic – the topic with the highest probability – for all tweets, then we calculated the log odds ratio of dominant topics among both groups to achieve a fair comparison, especially when there was a significant difference in number of tweets in each group. The log odds ratio of a topic can be interpreted as the probability of dominance of that topic in one group over another.

The odds that any topic T dominates in a group G are calculated as:

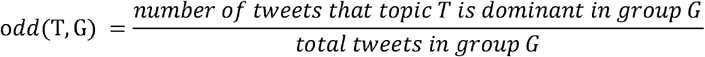

The log odds ratio of any topic T between two groups G_0_, G_1_ is calculated as:

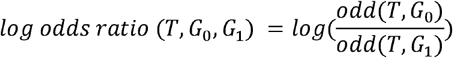

A positive log odds ratio indicates that topic T is more likely to appear in group G_0_, and a negative log odds ratio indicates that the topic T is more likely to appear in group G_1_. We did the same analysis to compare topic prevalence between hotspots and non-hotspots. We also implemented the chi-squared test and independent t-test to assess the differences in discussed topics across geographically grouped tweets. More specifically, the chi-squared test was used to determine whether there was a statistically significant difference between the expected dominant topic frequencies and observed dominant topic frequencies across the ADI groups and hotspot groups. We further leveraged independent t-tests to determine whether there was a difference between the means of the dominant topic probabilities in the low and high ADI groups.

## RESULTS

### Preprocessing and Integration of Tweets

Pre-processing resulted in 269,556 tweets (95.6% of the original geocoded dataset) from 119,611 Twitter users (out of which 63 users had more than 100 tweets). This dataset represents 1331 counties from all 50 states, the District of Columbia, and Puerto Rico. The range of the ADI is from 3 to 98. **Table 1** summarizes the characteristics of the final dataset.

**Table 1.**
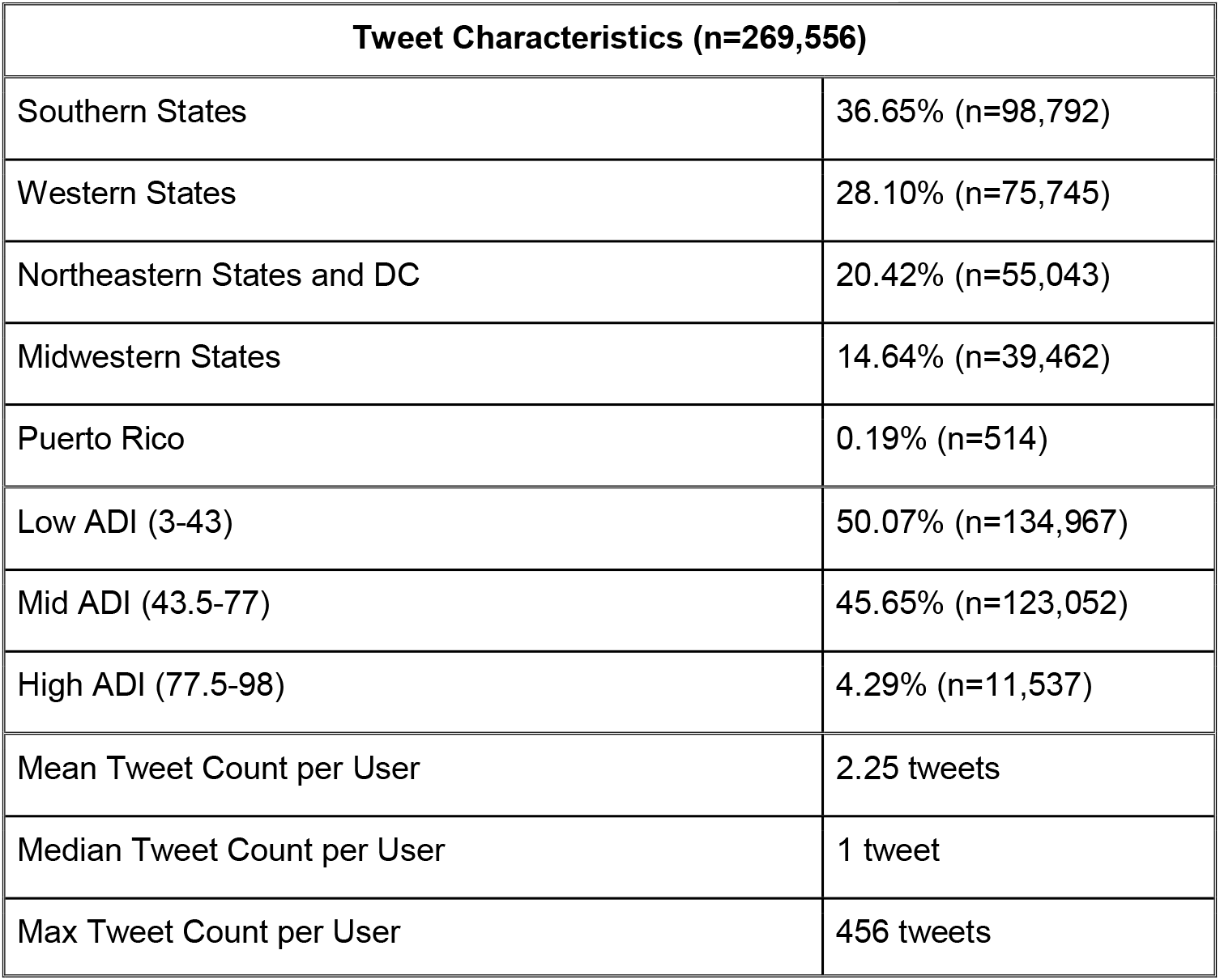
Characteristics of Dataset. Summary statistics of Twitter dataset in terms of user, geographic, and socioeconomic distribution.

### Topic Modeling

We evaluated models ranging from 10 to 50 topics and selected the model with 45 topics given that it had the highest coherence score (0.571). **Table 2** lists all 45 topics and **Table 3** presents examples of representative tweets (tweets with the highest probability for the given topic) for selected topics. Representative tweets for all topics are available in **Supplementary Table 1**. Topics were named based on the common theme of the top words. For example, we defined topic 1 as “Shopping” due to its top words “toilet”, “paper”, “store”, “shop”, “buy”, “walmart”, and “groceri” (stemmed version of groceries).

**Table 2.**
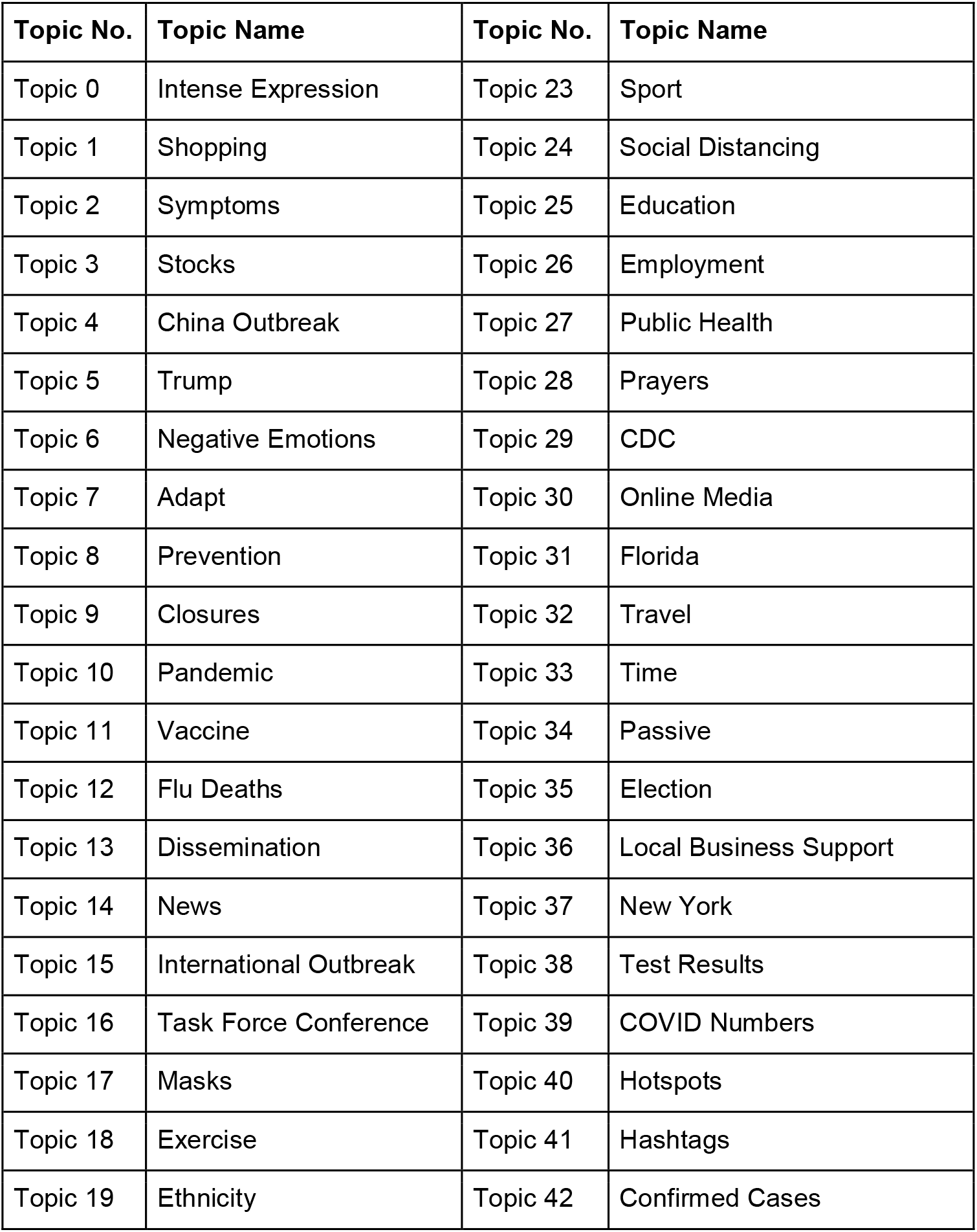
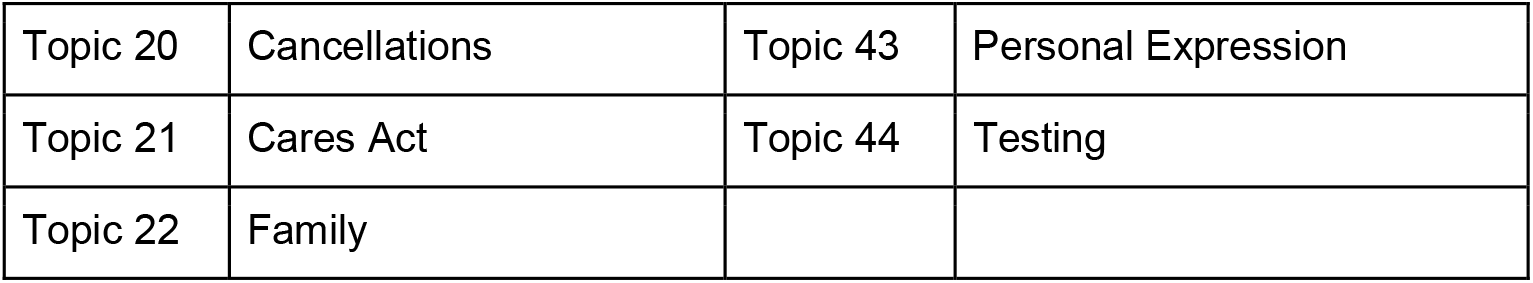
Identified topics based on LDA

**Table 3.**
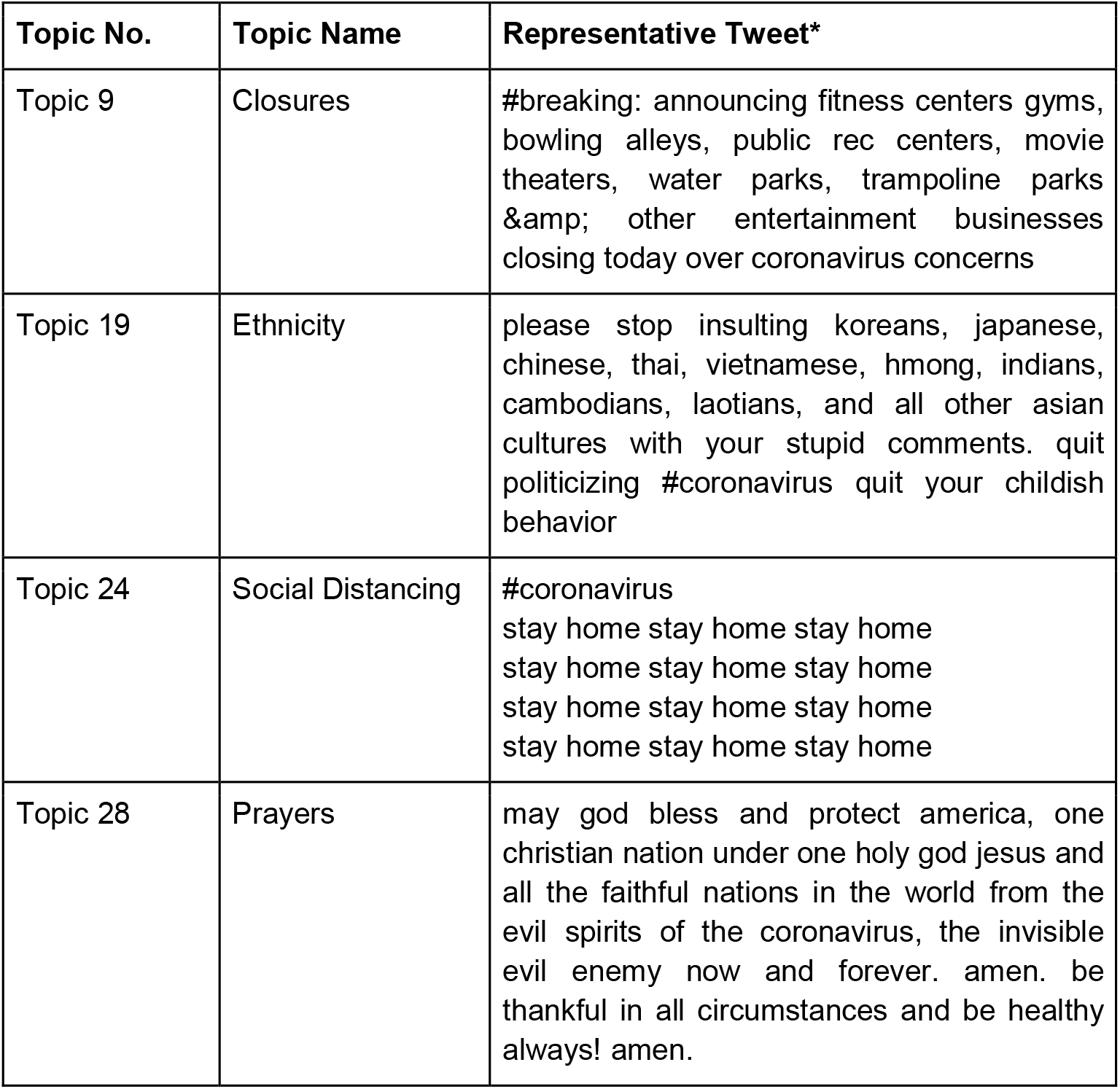
Example topics and the tweet with the highest probability of belonging to the topic. *Twitter handles removed to preserve Twitter users’ privacy without changing the meaning of the original tweets.

**In Figure 2**, we visualize a selected number of topics using word clouds. We show the top 10 words in each topic wherein the font size in each plot reflects the importance of a word in a specific topic. Word clouds for all topics are available in **Supplementary Figure 1**.

**Figure 2.**
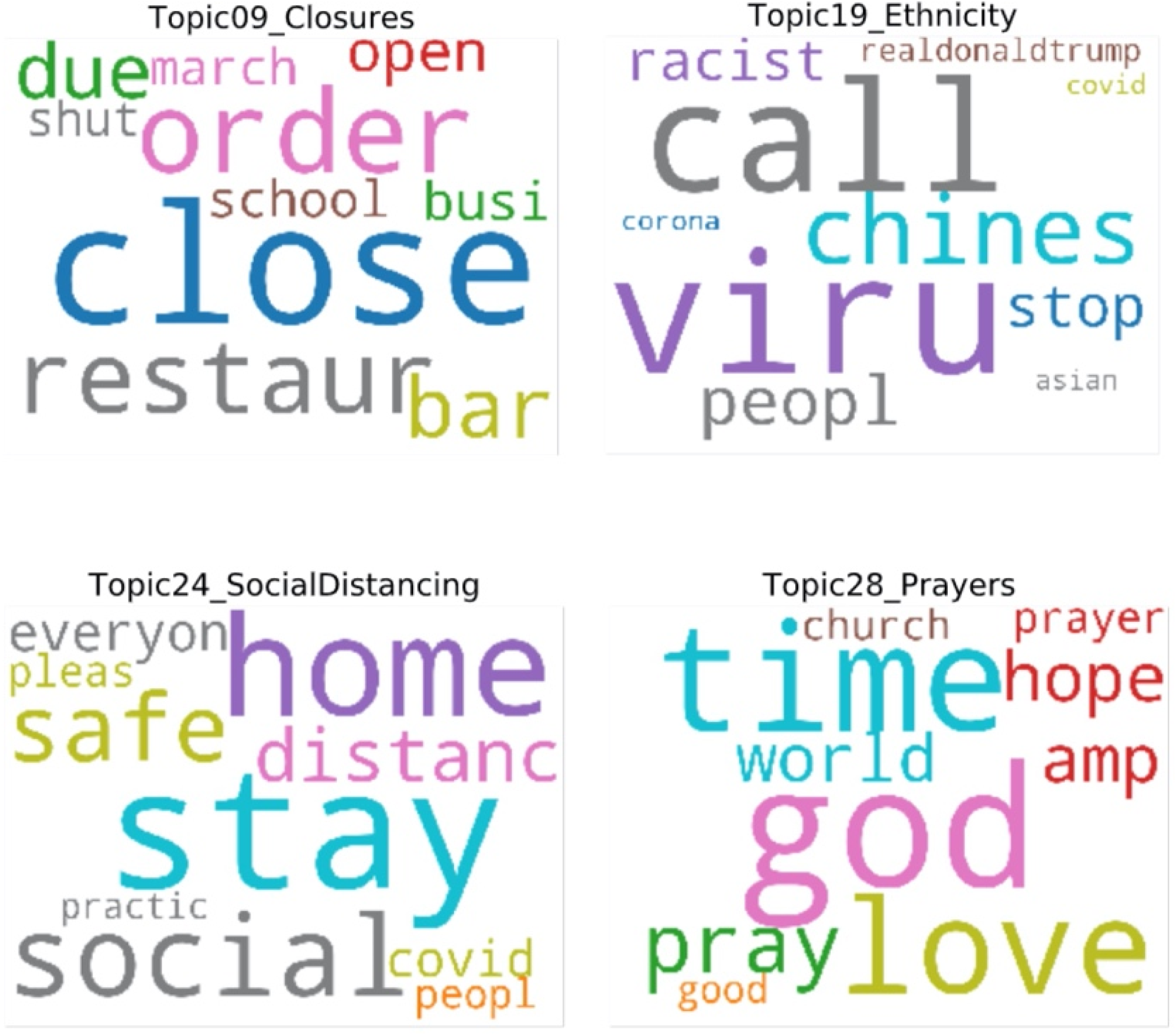
Visualization of the top 10 words in example topics.

### Comparing Topic Prevalence over Time

Through the monthly averaged distribution of topics, we delineated the topic dynamic from January to March and noted topics that peaked by month in **Figure 3**. For each month, topic prevalence compared to both of the other months had a significance of p <.0001 unless indicated otherwise. In January (**Figure 3A**), there were significant peaks in topics such as intense expression, negative expression, and personal expression (vs. Mar, p <.001) (Row 1). These topics are associated with profanity, anxiety, and emotions. We also noted a peak in discussion regarding an early understanding of the novel disease, namely symptoms, flu deaths, and preventative measures (vs. Feb, p <.01; vs. Mar, p <.05) (Row 2). Further, we noted significant discussion regarding China, international outbreak events (vs. Feb, p <.01; vs. Mar, p <.0001), and ethnicity (Row 3), as well as tweets concerning case counts (vs. Feb, p <.05; vs. Mar, p <.0001), hotspots (vs. Feb, ns; vs. Mar, p <.0001), and confirmed cases (Row 4).

**Figure 3.**
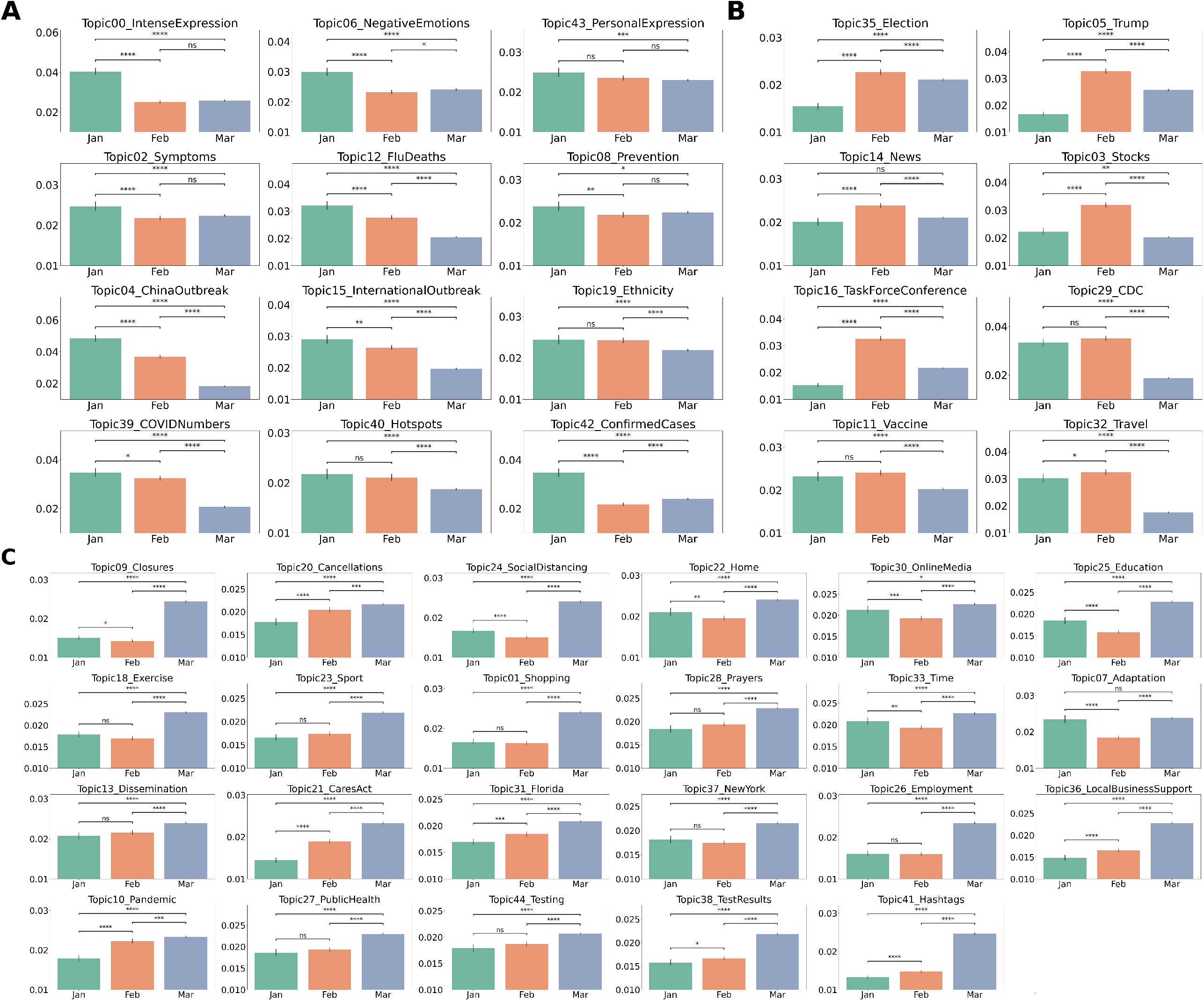
Distribution of topics grouped by month. A. Topics with higher proportions in tweets posted in January. B. Topics with higher proportions in tweets posted in February. C. Topics with higher proportions in tweets posted in March. Topics that had the same proportions for all months not shown. Significance testing results from two-sided Welch’s t-test with Bonferroni correction. Significance legend: ns: 5.00e-02 < p <= 1.00e+00. *: 1.00e-02 < p <= 5.00e-02. **: 1.00e-03 < p <= 1.00e-02. ***: 1.00e-04 < p <= 1.00e-03. ****: p <= 1.00e-04

In February (**Figure 3B)**, there was a significant rise in discussion surrounding the election, President Trump (Row 1), news articles, stocks (Row 2), the task force conference, and the CDC (Row 3). February also saw a significant discussion surrounding vaccines (vs. Mar, p <.0001) and travel (vs. Jan, p <.05; vs. Mar, p <.0001) (Row 4).

In March (**Figure 3C)**, there was a rise in discussions related to social distancing and disease mitigation strategies, namely closures, cancellations (vs. Jan, p <.0001; vs. Feb, p <.001), social distancing, staying home, online media (vs. Jan, p <.05; vs. Feb, p <.0001), and education (Row 1). In general, there were higher topic proportions of activities related to quarantine, in particular exercising, sport, shopping, prayers, words related to time, and adaptation (vs. Feb, p <.0001) (Row 2). March also resulted in more dissemination of information, discussion regarding the CARES Act, discussion of cases in Florida and New York, and tweets related to employment and local business support (Row 3). Finally, in March there was a significantly higher proportion of tweets related to the pandemic (vs. Jan, p <.0001; vs. Feb, p <.001), public health measures, tests and test results, and also a higher prevalence of COVID-related hashtags (Row 4).

### Comparing Topic Prevalence between Low and High ADI areas

ADI-specific analysis revealed significant differences in topic prevalence between low and high ADI areas. Comparing areas at the highest and lowest quintiles of ADI designation demonstrated differential effects (p <.001) in tweets by county level socioeconomic resourcing. Topics that are more likely to dominate in high ADI counties and low ADI counties are shown in **Figure 4A**. Tweets from high ADI areas are more likely to share emotional content with intense (p <.0001), negative (p <.01), personal expression (p <.01) or prayers (p <.05), as well as news regarding confirmed cases, the outbreak in China, flu deaths, and the CARES Act (all p <.0001). On the other hand, tweets from low ADI areas were more likely to discuss the impact of COVID-19 on hotspots, local businesses, and New York status (all p <.0001). Topics related to the larger public health crisis (p <.001) and pandemic (p =.001), as well as dissemination of information (p <.0001), stocks (p <.01), and the task force conference (p =.01), were also significantly more prevalent in tweets from lower ADI areas. These areas were also more concerned about the progress of potential treatments like vaccines (p <.001). While tweets with political topics about elections (p =.937) and president Trump (p =.605) were more likely to come from low ADI areas, the differences were not statistically significant. Observing the topic proportion progress from January through March (**Figure 4B**), we noticed that “Intense Expression” and “CARES Act” topics had consistent trends at both high and low ADI areas, with the high ADI areas having an overall higher daily average topic probability. Furthermore, topics associated with public health policies and disease mitigation strategies in March such as “Social Distancing” and “Local Business Support” arose in tweets from low ADI areas at a higher prevalence than tweets from high ADI areas.

**Figure 4.**
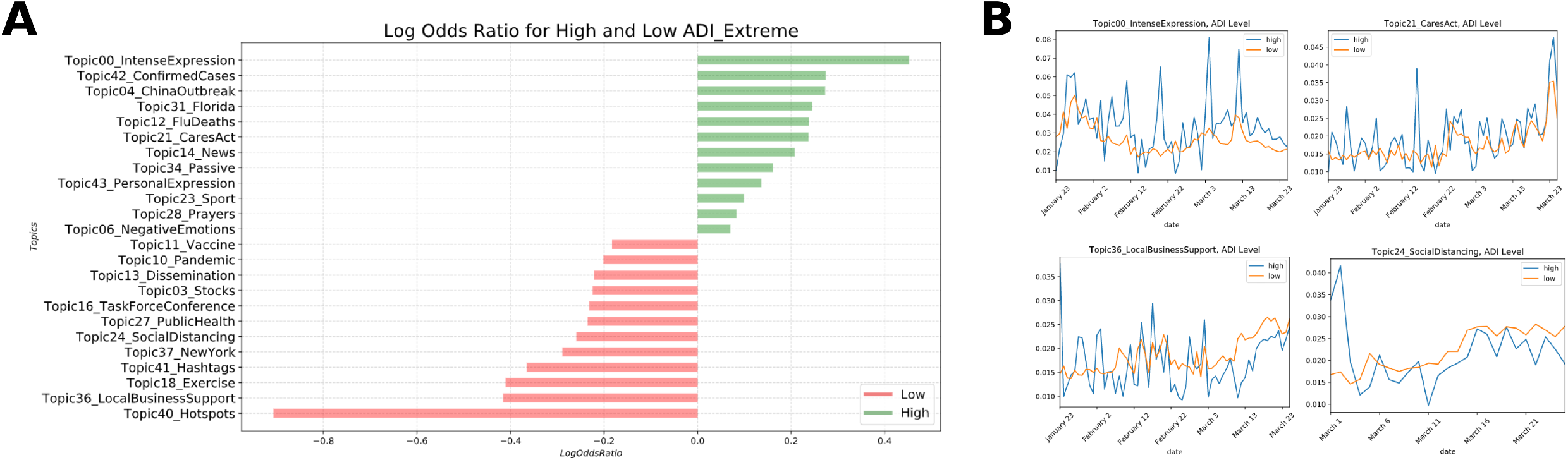
Topic prevalence comparisons between High and Low ADI based on Log odds ratio. A. Topics with significance difference between both groups (p <.05) B. Topic dynamic for example topics.

### Comparing Topic Prevalence between Hotspots and Non-Hotspots

The differences in the dominant topics’ prevalence between hotspots and non-hotspots areas were significant (p <.001). **Figure 5** demonstrates that tweets from hotspots had a higher log odds ratio for topics including New York, social distancing, public health and pandemic, information dissemination, exercise/sport, education, time, closures and employment. Tweets that were not posted from hotspots expressed negative or intense emotion, concern regarding the CDC guidelines and task force conference, international events and flu deaths, as well as stocks and shopping.

**Figure 5.**
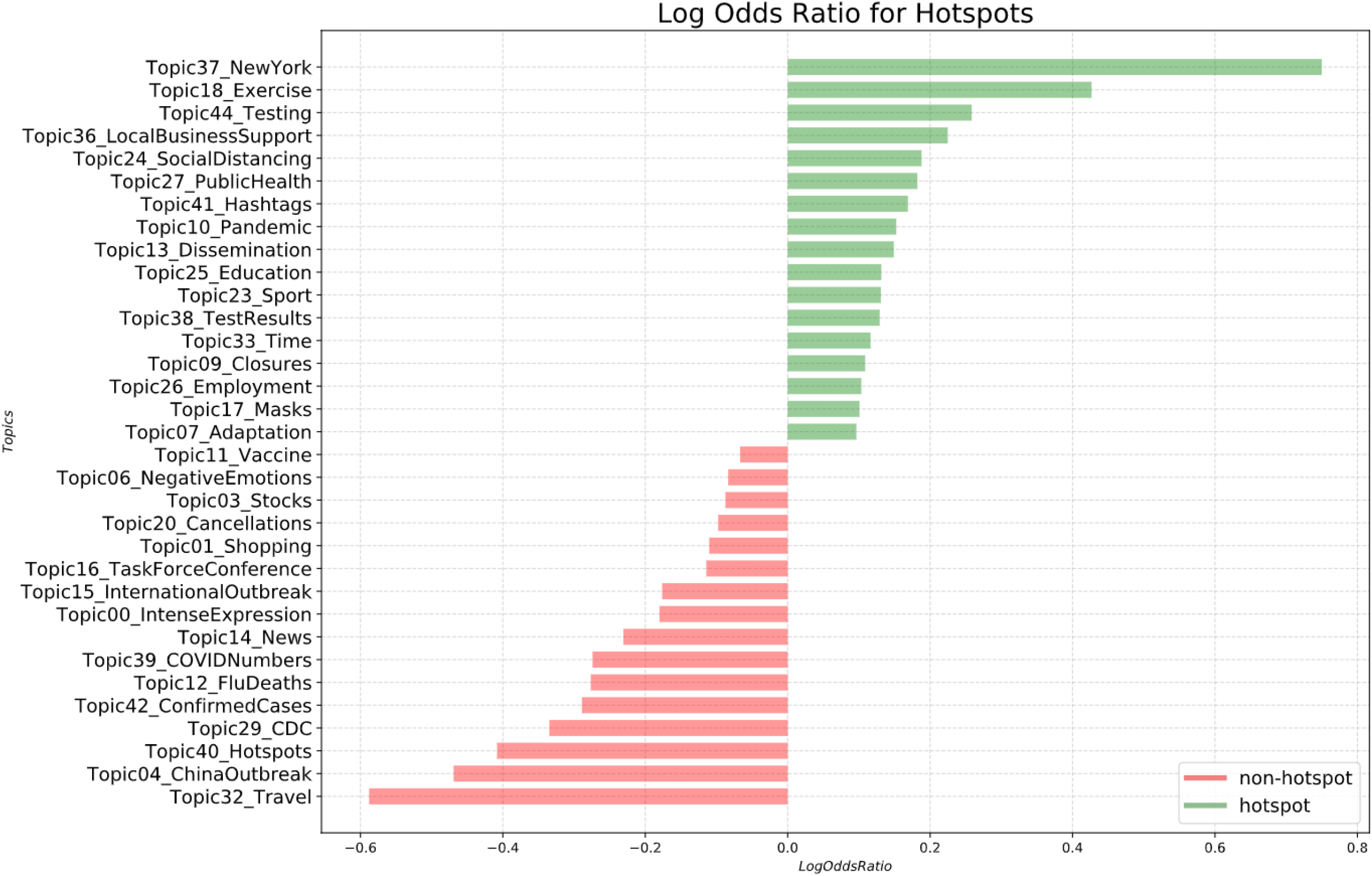
Topic prevalence between hotpots vs non-hotspots based on log odds ratio.

### Comparing Topic Prevalence Within Hotspots between Low and High ADI areas

We examined the tweets within the hotspots and compared topic prevalence between areas of high ADI and low ADI. **Figure 6A** demonstrates that tweets associated with confirmed cases, closures, intense expression, and hashtags were more prevalent from high ADI hotspots. Notably, tweets regarding employment concerns (p <.001) were also more likely to come from high ADI hotspots, which wasn’t significant in previous analysis comparing ADI and hotspots separately. Furthermore, tweets from low ADI hotspots were significantly more concerned with exercise, stocks, information dissemination, vaccine treatment, and cases in New York. We next observed the topic dynamics for selected topics from tweets collected in March (note that no high ADI areas were hotspots in January and February). There were notable spikes in employment concerns and intense expression from high ADI hotspots, whereas these topics remain consistent throughout the month for tweets from low ADI hotspots. Tweets about New York and social distancing remained consistently high in low ADI tweets.

**Figure 6.**
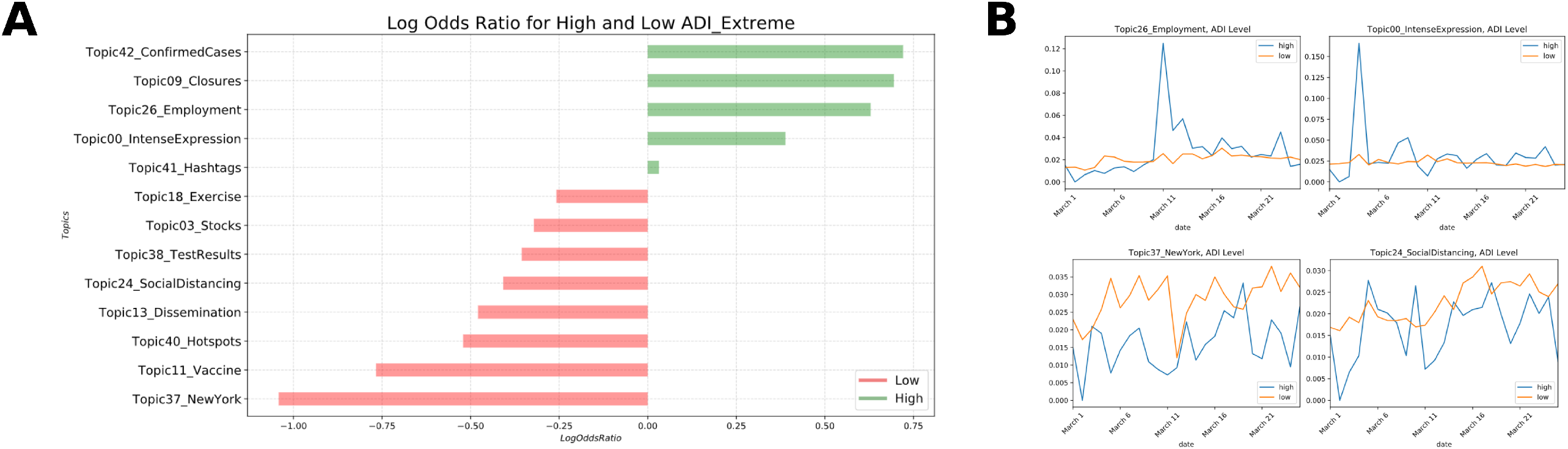
Topic prevalence comparisons within Hotspots between low and high ADI areas. A. Topics significant difference between the two groups (p <.05). B. Topic dynamic for example topics.

## DISCUSSION

Topic modeling of COVID-19-related social media data from Twitter demonstrates significant differences in individual responses to the pandemic based on geographic area, local disease prevalence and socioeconomic status. Over the progression of the crisis, tweets varied in topic, and these variations evolved over time and differed across counties with socioeconomic disparities. To our knowledge, this is the first study to link the ADI to geocoded tweets in order to explore the impact of geographic area-based socioeconomic status on tweet content.

This analysis follows the early pandemic timeline and establishes that topic modeling performs well in identifying major subjects of discussion on Twitter and successfully capture the nuances of their variability. Topic modeling has been applied to COVID-19-related tweets in an overlapping window of time (January 23 to March 7, 2020) [37], however limited topics, thus concerns, were identified and no analysis was reported about the emergence of topic during that period. As the first cases of COVID-19 broke news in January, we found the fear sentiment in tweets as people were broadly focused on disseminating as much information as possible. Similar conclusions were reported by Xue et. Al [37]. As time progressed, we found a massive increase in the number of tweets and there was increasing focus on local cases and events, public health information dissemination and testing, and quarantine activities.

Topic prevalence over time was explored in COVID-19 related tweets by Ordun et al [38], however, the analysis was limited to reporting trends and lacked extended investigations of linking the trending topics to other health or social factors. In our study, we have linked topic prevalence to socioeconomic status. Specifically, the topic prevalence comparisons between low and high ADI areas demonstrated that tweets from high ADI areas were more likely to share content regarding personal experiences, which ranged from positive affirmations of hope and prayers to negative or and intense expressions of anxiety or frustration. This was not surprising given that the disparate impact of the pandemic and the associated economic fallout have disproportionately impacted poorer communities, and Black and Hispanic communities have faced some of the highest rates of unemployment [39]. This was in many ways a result of the hard-hit industries overrepresented in these communities as well as inability to perform jobs in these industries from home [40, 41]. Furthermore, centuries of structural racism in the United States have led to lower resourcing in these areas and higher rates of medical co-morbidities that have been shown to increase COVID-19 risk [39] – all potentially contributing factors to an increase in intense, negative, and personal discussion in these areas pertaining to the public health and economic crisis.

Tweets from low ADI areas in March showed more discussion of social distancing and local business support, as quarantine policies hurt local businesses and resulted in discussions about bill relief to support these businesses. This result is consistent with the quicker response to stay at home orders from low ADI areas and is in line with recent reports of movement dynamic differences between low-income and high-income areas [42]. The higher prevalence of discussion surrounding stocks that was noted in low ADI areas was consistent with a greater stock market wealth residing amongst the wealthiest US households [43].

In the comparison between low and high ADI areas within hotspots, we identified that tweets with intense expression and those about employment insecurity were significantly more likely to come from high ADI hotspots, reinforcing the notion that, even after restricting to areas with high case counts, income and racial disparity result in disproportionate affects due to closures and job loss [41]. Furthermore, low ADI counties were significantly more concerned with information dissemination, cases in New York (on average a large low ADI hotspot), stocks, and vaccine treatment which follows our nationwide analysis of low ADI areas as showing increased focus on social and institutional reactions to the crisis.

Our approach of integrating a location-based socioeconomic index with Twitter topics offered increased insight into the topics inferred from the text, allowing a novel framework for assessing differential sentiment topics of conversation as they correlate to income, education, and housing disparities. Our integration of published COVID-19 hotspots further enables time-specific information of disease spread and how this corresponds to topics discussed on Twitter. These nuances are valuable for recognizing how public health communication, resource allocation policy, and information dissemination can best shape health crisis response to the needs of different communities, especially those with the lowest health resourcing, in future waves of the pandemic and emerging infectious disease outbreaks. Future public health efforts may use Twitter topic modeling to target messaging to the unique concerns of local communities and study the impact of health resource utilization.

### Limitations

Our study successfully explored on the pandemic topics of conversation across tweets. However, there were a few limitations. For technical reasons on the server, fewer tweets were scraped on some dates. However, our previous work on [44] has shown that that we were still able to glean valuable conclusions from our data that represent the early pandemic progression. Another limitation for all Twitter-based research is that tweets posted from private accounts could not be retrieved from the API. Furthermore, due to restrictions with Twitter geocoding, there was some degree of positional inaccuracy that we accepted in our study design in that we were only able to collect geographic coordinates to the resolution of a county, and therefore characterized each tweet by the county rather than the census tract or block group. Given the inherent geographic masking techniques used by Twitter to promote confidentiality, and our study design which involved cross-area estimation and simple geographic centroid assessment [19], we acknowledge aggregation bias as a study limitation. Despite this, however, we found that, on average, the county ADI was distributed such that the median ADI was a reasonable approximation for the county.

## CONCLUSION

Twitter analysis linking geocoded tweets to markers of geographic socioeconomic resourcing demonstrates that the COVID-19 pandemic has differentially impacted areas of the United States that are already institutionally underserved. This finding highlights the need to address the specific fears and concerns of these communities through personalized public health messaging and policy reform, addressing consistent issues such as job security and negative emotions likely associated with greater instability during the crisis. Our work indicates the emerging utility for linking natural language processing techniques to analyze real-time social media data and measures of social determinants of health.

## Data Availability

Publicly available Tweets downloaded using the Twitter API.

## CONFLICT OF INTEREST STMT

There is no conflict of interest.

## FIGURE LIST

Supplementary Figure 1. Visualization of the top 10 words in all topics.

