## Supplementary figures and images for "COVID-19 Twitter-based analysis reveals differential concerns across areas with socioeconomic disparities"

### Supplementary Figure 1

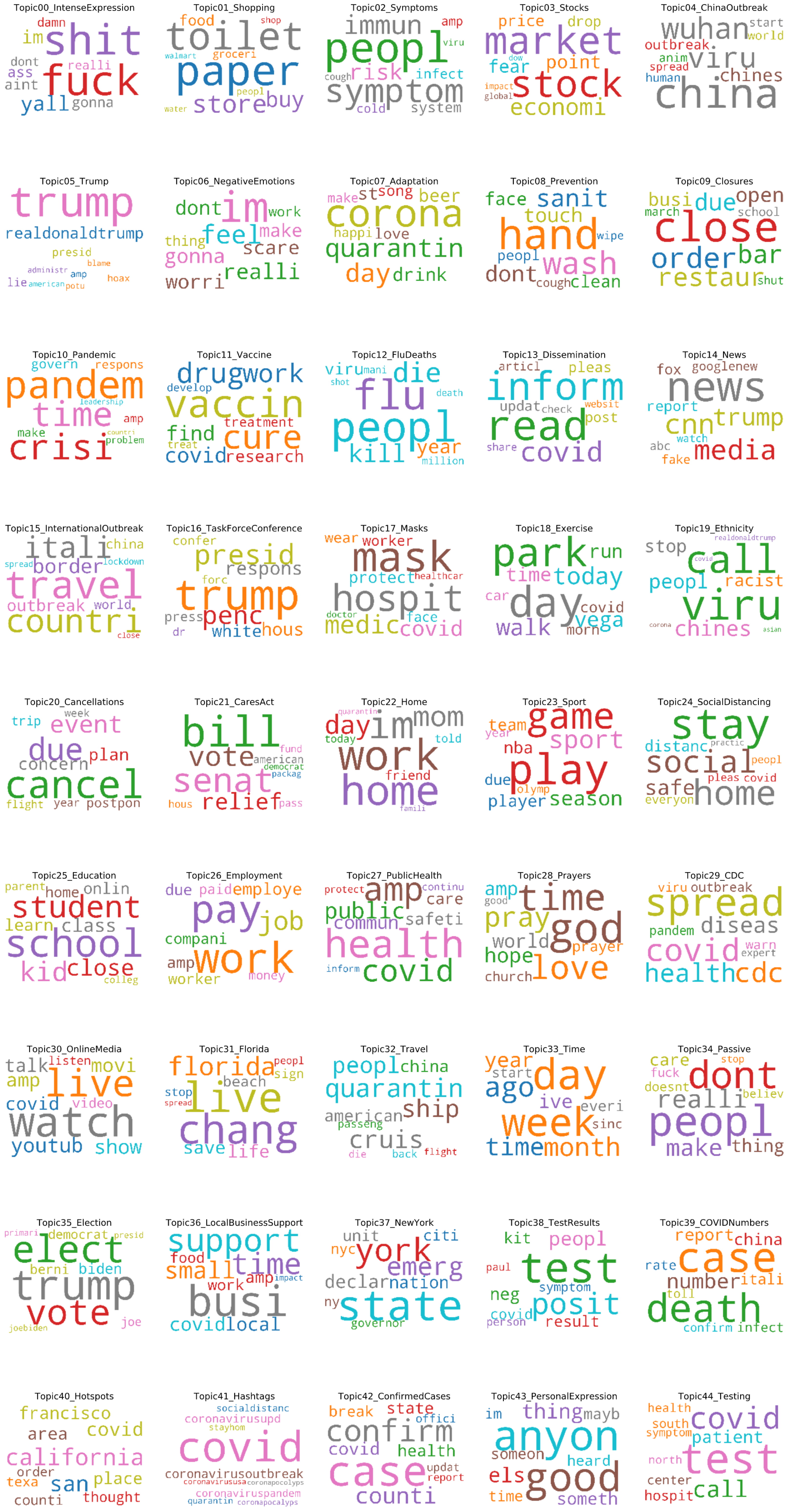
