## Supplementary Table 1 for "COVID-19 Twitter-based analysis reveals differential concerns across areas with socioeconomic disparities"

Supplementary Table 1. All topics and representative tweets. Example topics and the tweet with the highest probability of belonging to the topic. *Twitter handles removed to preserve Twitter users’ privacy without changing the meaning of the original tweets.

| **Topic No.** | **Topic Name** | **Representative Tweet*** |
| --- | --- | --- |
| Topic 0 | Intense Expression | this is for the coronavirus you big fat white nasty smellin fat bitch why you took me off the mf schedule with yo trifflin dirty white racist ass big fat bitch oopa loopa body ass bitch im comin up there and im gon beat the fuck outta you bitch |
| Topic 1 | Shopping | [redacted for privacy] |
| Topic 2 | Symptoms | coronavirus symptoms  fever, cough, shortness of breath  allergy symptoms  itchy eyes, stuffy nose, sneezing  influenza symptoms  fever, cough, body aches, fatigue, chills, headache, possibly sneezing, stuffy nose &amp; sore throat  reminding to know the difference |
| Topic 3 | Stocks | according to stock sales disclosures by senators after a closed door briefing on january 24 about the coronavirus threat, the following senators sold stocks:  senator richard burr  senator kelly loefner  senator dianne feinstein,  sena |
| Topic 4 | China Outbreak | the world s only scaly mammal, the pangolin, has been linked by chinese scientists to the spread of the wuhan coronavirus, widely trafficked as a source of both meat and of scales for use in traditional medicine, and may be the intermediate host between bats and humans. |
| Topic 5 | Trump | biden campaign accuses trump of trying to 'rewrite history' on his 'failed' coronavirus leadership #topbuzz **thanks for exposing lying unfit putin puppet trump and his swamp administration. |
| Topic 6 | Negative Emotions | i make jokes about the coronavirus because i actually have really intense fears and anxiety over this kinda stuff and if i don t make jokes i ll probably lose my mind |
| Topic 7 | Adaption | #jamiphy  ig: leahmurphymusic  .  .  .  #music #musician #musicians #guitar #guitarist #piano #pianist #bass #bassguitar #bassist #drums #drummer #sing #singer #rap #rapper #rock #hiphop #pop #blues #jazz #concert #band #dance #art #love #beautiful #coronavirus #covid19 |
| Topic 8 | Prevention | wash your hands! wash your hands! wash your hands! wash your hands! wash your hands! wash your hands! wash your hands! wash your hands! wash your hands! wash your hands! wash your hands! wash your hands! wash your hands! #coronavirus #covid 19 |
| Topic 9 | Closures | #breaking: @govmikedewine announcing fitness centers gyms, bowling alleys, public rec centers, movie theaters, water parks, trampoline parks &amp; other entertainment businesses closing today over coronavirus concerns |
| Topic 10 | Pandemic | one of the saddest aspects of this tragedy is that it s not a national conversation right now. gun violence isn t unique enough... especially with the current coronavirus obsession. we can prevent these situations, there are actions we can take. this is devestating; it has to |
| Topic 11 | Vaccine | french peer-reviewed study: our treatment cured 100% of coronavirus patients.  anti-malaria drugs already developed have killed 100% of test-tube coronavirus being reported around the world  amazing work being done |
| Topic 12 | Flu Deaths | #coronavirus kills young people #farmers #education #teaparty #aarp #veterans #metoo #genx #millenials #marchforourlives #1u #maga #independents #indigenous #p2 #latinx #blm #lgbtq |
| Topic 13 | Dissemination | additionally, there is a faq page that is being updated frequently as more information comes in. go to suggest an faq to submit more questions you might have so we can better track and respond to questions, which are many at this time.  faq page: |
| Topic 14 | News | msnbc s rachel maddow issued an impassioned plea for news networks to stop broadcasting donald trump s daily press briefings about the coronavirus pandemic on live tv. # via |
| Topic 15 | International Outbreak | venice and milan among major italian cities locked down as 16million put in quarantine  the italian government has quarantined the entire region of lombardy and neighbouring areas, including venice and milan, to halt the spread of coronavirus |
| Topic 16 | Task Force Conference | "in 2018, the trump administration fired the government s entire pandemic response chain of command, including the white house management infrastructure. " |
| Topic 17 | Masks | if you hoarded face masks, gloves and other things that medical professionals need more than you do right now, please find a way to safely donate them. their lives &amp; others may depend on it.  doctors say shortage of protective gear is dire |
| Topic 18 | Exercise | overwhelmed today. went on a nice, eerily quiet walk in the snow with ocean this evening, it was beautiful. one day at a time.  .  .  .  #quarantine #denver #colorado #snow #snowing #snowday #coronavirus #walk |
| Topic 19 | Ethnicity | please stop insulting koreans, japanese, chinese, thai, vietnamese, hmong, indians, cambodians, laotians, and all other asian cultures with your stupid comments. quit politicizing #coronavirus quit your childish behavior |
| Topic 20 | Cancellations | ultra s march festival canceled over #coronavirus fears in #miami #sxsw v #f1 #motogp |
| Topic 21 | Cares Act | chuck schumer, dems demand senate immediately pass pork-filled coronavirus bill 'as-is' now is the time for to take head-on and tell america about all dem pork in bill, like $1b for abortions!!! via |
| Topic 22 | Home | welcome to marriage during #coronavirus #quarantine  day 4 hubby has been off work and home    husband: baby i m putting my headsets on to get some work done  me: okay  me: start talking to hubby 5 minutes later. lol  him: baby(stares) i can t hear you |
| Topic 23 | Sport | hey kshsaa give these seniors who play spring sports 2 months of eligibility to get to play their spring sports due to this coronavirus pandemic they deserve to play senior year sports so give them another year they deserve to play their senior y |
| Topic 24 | Social Distancing | #coronavirus  stay home stay home stay home  stay home stay home stay home  stay home stay home stay home  stay home stay home stay home |
| Topic 25 | Education | #covid19 update: cusd schools will close effective mar. 16 apr. 3. spring break is now scheduled next week mar. 16-20, with school closed an additional 2 weeks through apr. 3. review the complete announcement to families + next steps on #cusdinsider: |
| Topic 26 | Employment | 90- day delay in tax payments deadline!  treasury secretary steven mnuchin announced that individuals and corporations can delay their tax payments for 90 days from the april 15 deadline.  during that time, the irs will not charge interest or penalties. |
| Topic 27 | Public Health | nonprofits like the ymca are serving #pennsylvania communities during the coronavirus crisis, but the economic downturn makes it harder to provide emergency child care and feed kids. please support $60b in support for nonprofits! #relief4ch |
| Topic 28 | Prayers | may god bless and protect america, one christian nation under one holy god jesus and all the faithful nations in the world from the evil spirits of the coronavirus, the invisible evil enemy now and forever. amen. be thankful in all circumstances and be healthy always! amen. |
| Topic 29 | CDC | so. it. has. begun. #2019coronavirus #coronavirus #2019ncov #2019n_cov #coronaoutbreak #virus_corona #virusoutbreak #ncov2019 #chinaoutbreak #chinapneumonia #china #wuhanpnemonia #wuhanoutbreak #ncov19 #broitsjusttheflu |
| Topic 30 | Online Media | i am supporting @elijahdaniel's #cultforgood project to bring hundreds of thousands of necessities and free coronavirus testing to the homeless population during this covid-19 outbreak, we still need help  hmu if u can sponsor or donate product!!! |
| Topic 31 | Florida | mayor bill de blasio: close nyc public schools to slow the spread of coronavirus - sign the petition! via |
| Topic 32 | Travel | just in: flight from wuhan, china, has landed at march air reserve base in riverside 200+ american citizens onboard will be quarantined for 72 hours and screened for #coronavirus symptoms. #abc7eyewitness |
| Topic 33 | Time | all us citizens are entitled to 700 usd per week to stay at home to avoid the spread of covid-19 coronavirus starting from march 23,2020 the government grant pay is accessible to all no matter employment status. read full article here on how to claim  ... |
| Topic 34 | Passive | the most annoying thing is when people who clearly understand absolutely nothing about coronavirus tell me that im being ridiculous for telling people that its serious |
| Topic 35 | Election | trump is attacking joe biden.  bernie is attacking joe biden.  meanwhile, joe biden is attacking coronavirus.  and that s the difference. |
| Topic 36 | Local Business Support | as part of the federal relief package, the federal sba will be providing disaster assistance to small businesses affected by the covid-19 pandemic. the program is intended to provide low-interest working capital to small business. #coronavirus #covid19 |
| Topic 37 | New York | washington state gov. jay inslee issues stay-at-home order amid the coronavirus outbreak - |
| Topic 38 | Test Results | although, he s feeling fine &amp; is asympomatic... rand paul gets tested &amp; becomes the first us senator to test positive for #coronavirus  so... maybe we need to be testing everyone, not just rich politicians &amp; pro athletes! |
| Topic 39 | COVID Numbers | #coronavirus mortality rate in #italy now 3.4pct...putting it on par with the reported mortality rate in hubei. 11 dead out of 323 cases. not sure yet but think italy has surpassed south korea death toll. iran no. 1 after china. |
| Topic 40 | Hotspots | your life album is out  #newmusic #music #newalbum #album #piano #vocal #guitar #synth #beat #mixtape #lit #hype #fire #musicproducer #singersongwriter #songwriter #producer #record #recording #time #azmusic #arizona #phoenix #love #coronavirus |
| Topic 41 | Hashtags | amen! #covid 19uk #covidontario #highriskcovid19 #democraticdebate #safeathome #covid 19 #covid 19 #coronavirusupdates #coronavirus #coronavirusoutbreak #coronaindia #covid19 #mondaymotivation #mondaymood #mondaythoughts #seattle #seattlecoronavirus #seattlecovid19 |
| Topic 42 | Confirmed Cases | tuesday coronavirus update:  564 confirmed cases in ohio  11 confirmed cases in lucas county  2 confirmed cases in defiance county  1 confirmed case in sandusky county  2 confirmed cases in wood county  8 deaths in ohio  1 death in lucas county |
| Topic 43 | Personal Expression | with everything that s going on, i figured i d talk about novel coronavirus-19 (as if you don t hear enough already).i hope what i ve written helps clear whatever weird things you ve heard in the media because it s |
| Topic 44 | Testing | 2/ by phone: the lawrence general hospital covid-19 (coronavirus 2019) community screening line is staffed by one of our nurses 24 hours a day, 7 days a week and can be reached by calling us at 978-946-8409. |
